# Fatigue presentation, severity, and related outcomes 12 weeks post-COVID-19 hospitalization

**DOI:** 10.1101/2022.10.18.22280199

**Authors:** Tianna Magel, Emily Meagher, Travis Boulter, Arianne Albert, Melody Tsai, Carola Muñoz, Chris Carlsten, James Johnston, Alyson Wong, Aditi Shah, Chris Ryerson, Rhonda Jane Mckay, Luis Nacul

**Affiliations:** Women’s Health Research Institute, British Columbia Women’s Hospital, Vancouver, BC, Canada V6H 3N1; Division of Respiratory Medicine, Department of Medicine, University of British Columbia, Vancouver, BC, Canada V5Z 1M9; General Internal Medicine, Department of Medicine, University of British Columbia, Vancouver, BC, Canada V6Z 1Y6; Faculty of Medicine, Department of Family Practice, University of British Columbia, Vancouver, BC, Canada V6T 1Z3

## Abstract

**Background:** Increasing evidence on long-term health outcomes following SARS CoV-2 infection shows post-viral symptoms can persist for months. The aim of the present study was to examine the prevalence and outcome predictors of post-viral fatigue and related symptoms 3 and 6 months following infection.

**Methods:** A prospective cohort of patients hospitalized with COVID-19 (n=88) were recruited from a Post-COVID-19 Respiratory Clinic (PCRC) in Vancouver, Canada to examine predictors of long-term fatigue and substantial fatigue. Multivariable logistic and linear regression analysis were used to examine the relationship between patient predictors (medical history at the time of hospitalization and follow-up patient-reported outcome measures) and the presentation of fatigue and substantial fatigue at 3 and 6 months follow-up.

**Results:** The number of patients exhibiting fatigue and substantial fatigue was 58 (67%) and 14 (16%) at 3 months and 47 (60%) and 6 (7%) at 6 months post-infection, respectively. Adjusted analysis revealed the number of pre-existing comorbidities to be associated with fatigue (OR 2.21; 95% CI 1.09-4.49; 0.028) and substantial fatigue (OR 1.73; 95% CI 1.06-2.95; 0.033) at 3 months follow-up. Except for shortness of breath, self-care, and time, all follow-up variables were found to be associated with fatigue and substantial fatigue at 3 months follow-up.

**Conclusion:** Fatigue and substantial fatigue are common, and decrease from 3 to 6 months; however, a significant number of patients continue to exhibit long-term fatigue at 6 months follow-up. Further research is needed to clarify the causality of viral infections and co-factors in the development and severity of fatigue as a symptom and in meeting post-viral fatigue syndrome or ME/CFS diagnostic criteria.

## Introduction

Rapid global spread of COVID-19 has resulted in an estimated 621 million confirmed cases and approximately 6 million deaths worldwide as of October 2022.^1^ There have been extensive investigations into acute stages of viral infection however, less is known about the long-term impacts experienced after infection. As the number of patients who have recovered from COVID-19 grows, it is evident that “recovery” is not synonymous with a return to previous health status for many individuals.

Emerging evidence demonstrates that, for a significant number of individuals, post-COVID-19 sequalae persists well beyond the acute stages of viral infection.^2–5^ Estimates for the proportion of people who experience post-COVID conditions vary. However, symptoms that persist for more than 12 weeks are termed “Long-COVID’ or Post-Acute Sequalae of SARS-CoV-2. ^6,7^ The CDC estimates that over 30% of hospitalized individuals experience post-COVID related symptoms for six months or longer after infection.^8^

COVID-19 is now recognized as a systemic disease with multiorgan involvement.^9^Fatigue and cognitive impairment, along with abnormal respiratory function and other enduring neuropsychiatric and physical manifestations have been reported as the most prevalent and debilitating symptoms of post-COVID conditions.^3,10,11^ Literature examining long-term outcomes shows, for those hospitalized, post-COVID-19 symptoms can persist for upwards of 12 months.^4^ Indeed, many may experience long-term symptoms congruent with that of Myalgic Encephalomyelitis /Chronic Fatigue Syndrome (ME/CFS), such as persistent or debilitating fatigue, also referred to as post-COVID-19 fatigue syndrome.^3,7^

Fatigue and other long-COVID related outcomes are varied across studies due to poor standardization of data collection methods and measurement tools.^12^ Standardized investigations into the presentation of fatigue and related symptoms after viral infection is critical to providing evidence-based post-viral care for those experiencing severe long-term forms of COVID-19 infection. In this paper, we examine the clinical presentation of post-viral fatigue within a prospective cohort of individuals hospitalized with severe COVID-19 in Vancouver, Canada.

Hospitalization and follow-up predictors of fatigue and patient-reported clinical outcomes are described at 3- and 6-months post-symptom onset of SARS-CoV-2 infection.

## Methods

### Measurements

This study involved a prospective consecutive cohort of 88 individuals ≥18 years recruited from the Post-COVID-19 Respiratory Clinic (PCRC) in Vancouver, Canada. Participants included those with a confirmed SARS-CoV-2 infection who were hospitalized from March to June 2020. Detailed methods, 3- and 6-month respiratory outcomes, and some patient-reported outcome measures (PROMs) have been previously reported elsewhere.^13,14^ Briefly, at hospitalization and 3- and 6-months post-infection, patient medical history, clinical variables, and patient-completed standardized questionnaires were collected. Clinical predictors of fatigue included age, sex, number of pre-existing comorbidities, ICU admission, and mechanical ventilation. Patient-reported predictors, collected at 3- and 6-months, included scoring on the EuroQoL 5-Dimensions (EQ-5D), the Frailty Index (FI), the University of California San Diego Shortness of Breath Questionnaire (UCSD), Patient Health Questionnaire-9 (PHQ-9), and the Pittsburgh Sleep Quality Index (PSQI). The EQ-5D is a five dimension preference based quality of life assessment. Scores from the EQ-5D were converted to a health utility index whereby scores of 1 represent perfect health and scoring of 0 is death.^15^ The UCSD was used to assess patient dyspnea with scores greater than 10 reflecting dyspnea.^16,17^ PHQ-9 scores assessed the presence and frequency of depression.^18^ Assessments of sleep quality were ascertained using the PSQI and aging and vulnerability to adverse outcomes were determined using the FI.^19,20^

### Outcomes

Primary outcomes included the presentation and severity of persistent fatigue at 3- and 6-months post-symptom onset. Fatigue was classified as an individual reporting feeling tired or having little energy for several days or more over the last two weeks (PHQ-9) as well as, indicating “always feeling tired” (FI). Substantial fatigue was classified as an individual reporting fatigue, as defined above, and “slight” differences in their ability to conduct usual activities (EQ-5D).

### Statistical Analyses

Descriptive statistics were used to describe participant characteristics at the time of hospitalization. Associations between all predictors (i.e. clinical and PROMs) and presentation of fatigue and substantial fatigue at 3- and 6-months were examined using multivariable logistic regression modelling. Adjusted analysis of all predictor variables at 3- and 6-month timepoints did not include the EQ-5Ds “self-care” dimension due to a lack of variance in responses. Given the low number of patients with substantial fatigue at 6 months follow-up (n= 6), modelling for substantial fatigue included only the 3-month follow-up period.

McNemar’s test was then used to examine changes in the presence of substantial fatigue across time. Relationships between clinical predictors and PROMs were examined using multivariable linear regression. Statistical significance was determined by a two-sided p-value of <0.05. Analyses were conducted using the statistical software package R (Version 4.2.1).

## Results

Table 1 shows the main characteristics of the cohort at hospitalization and 3- and 6-months as well as the prevalence of fatigue at 3- and 6-months. Among those included in this analysis (n=88), mean age was 61.1 (±16.2) years; 63.6% were male, 45.5% identified as white; 80.7% and 89.8% had no pre-existing lung or autoimmune disease, respectively. Approximately 48.2% were admitted to ICU, of whom 20.5% were on mechanical ventilation during hospitalization. Prevalence of fatigue and substantial fatigue were reported to be 66.7% and 16.1%, respectively, at 3 months. By 6-month follow-up, fatigue was exhibited in 59.5% and substantial fatigue in 6.9% of patients.

**Table 1.**
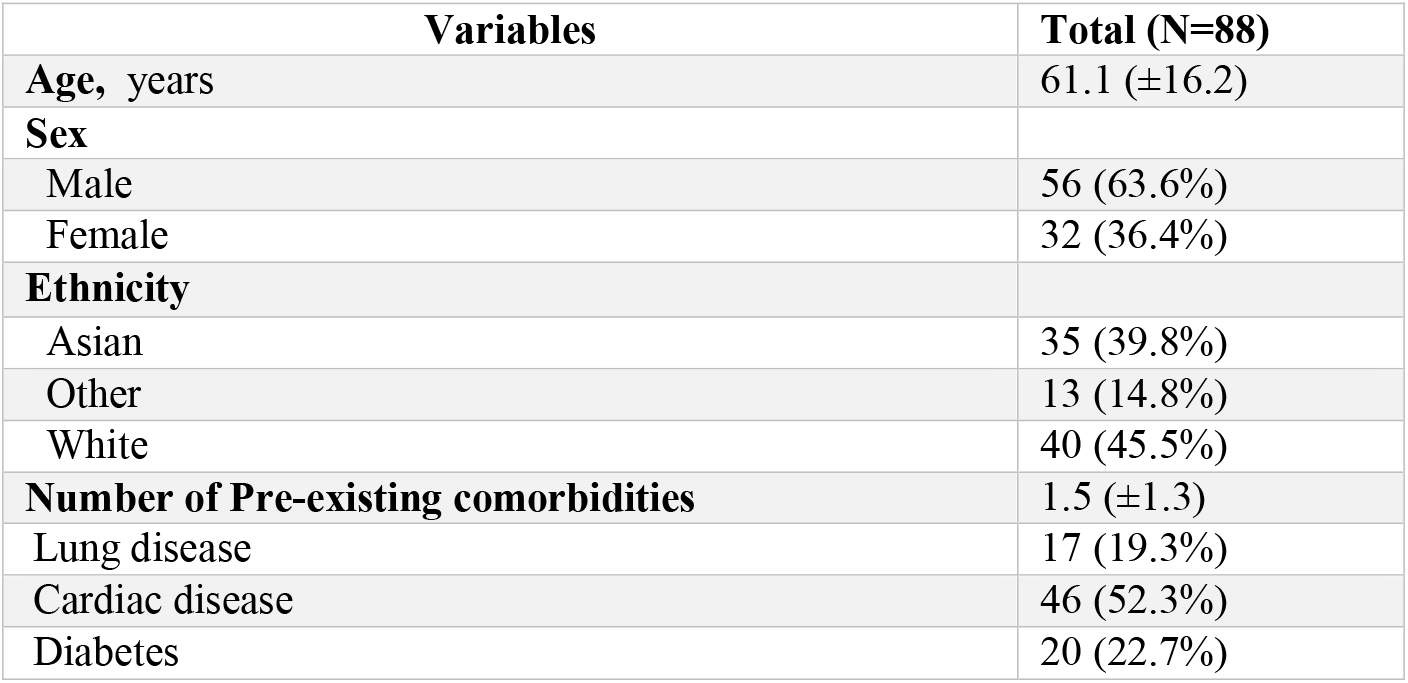
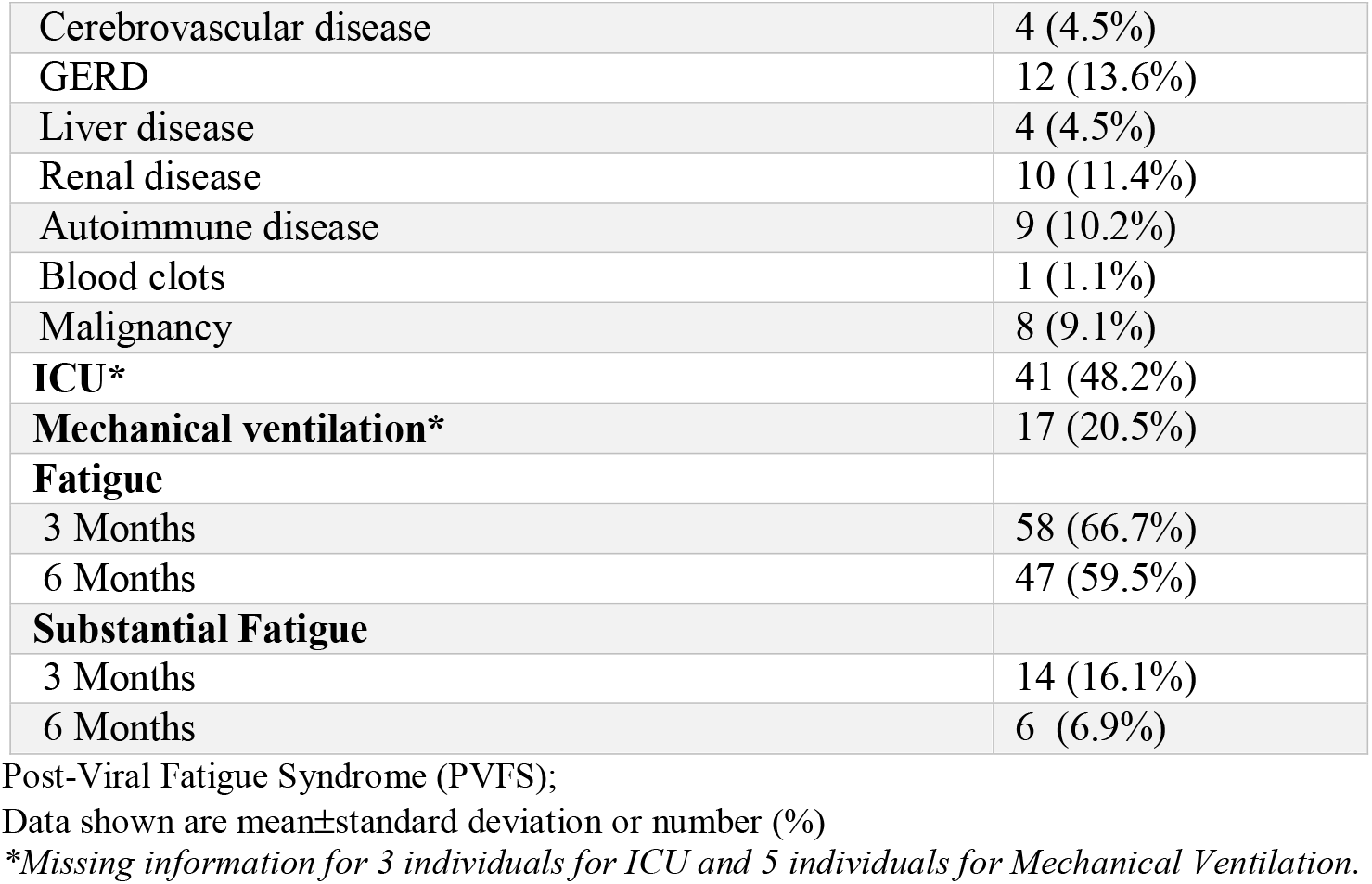
*Characteristics of PVFS cohort* and presence of Fatigue at 3- and 6-months.

### Hospitalization Predictors of Fatigue at 3 and 6 months

The number of pre-existing comorbidities had a trend toward association with fatigue at 3 months (p = 0.06). Adjusted multivariable analysis, controlling for other variables at the time of hospitalization, revealed age (OR 0.93; 95% CI 0.88-0.98; p = 0.012) and number of comorbidities (OR 2.21; 95% CI 1.09-4.49; p = 0.028) were associated with fatigue at 3 months post-viral infection (*Figure 1*). Patients who did not exhibit fatigue at 3 months did not go on to exhibit fatigue at 6 months follow-up.

**Figure 1.**
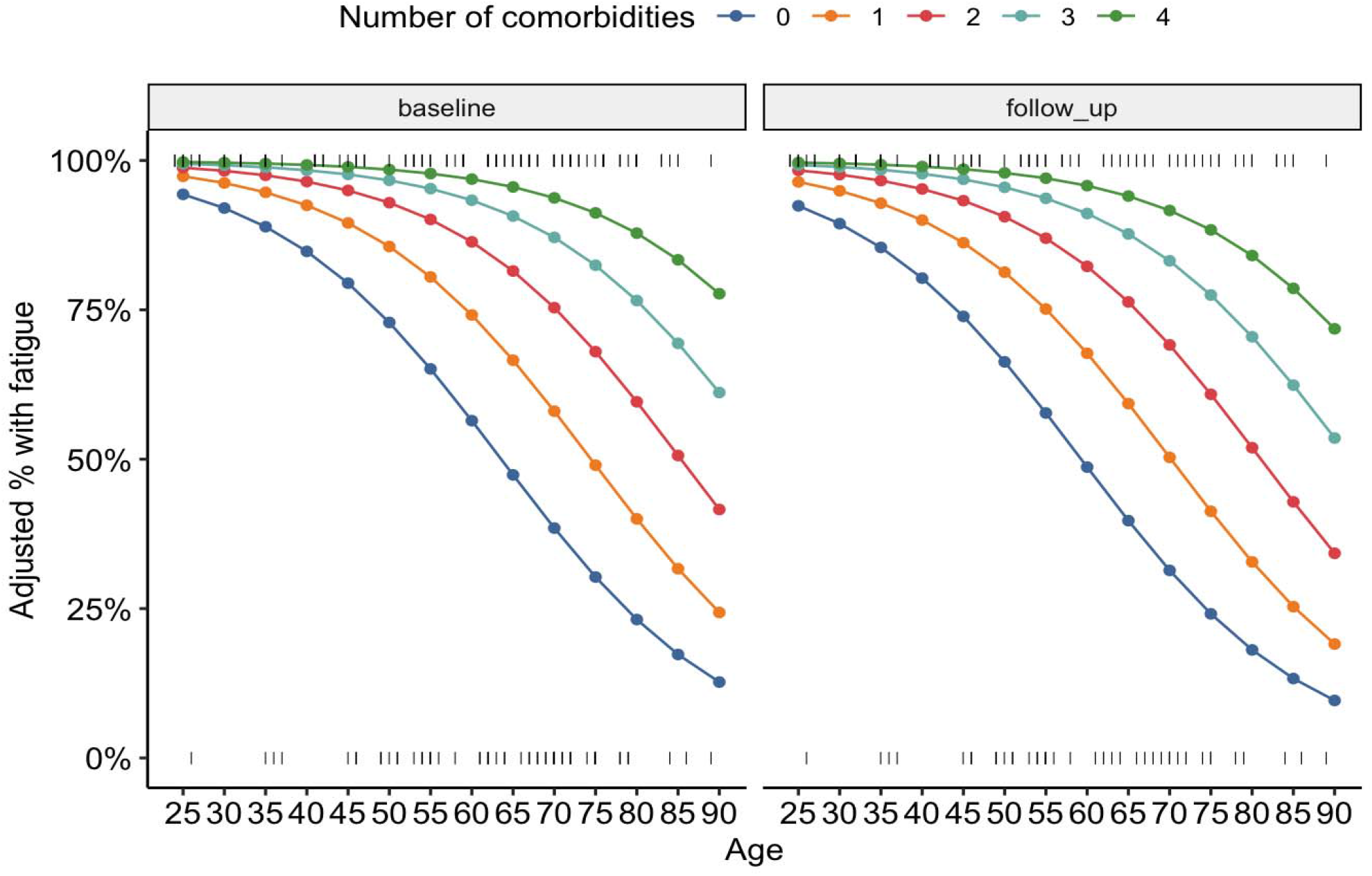
Fatigue and number of comorbidities at hospitalization and follow-up. The present figure shows the relationship between age, comorbidities and proportion with fatigue controlling for other variables in the multivariable model (no difference in proportions at 3 and 6 months).

### Predictors of Fatigue at 3 and 6 months

Adjusted logistic regression analysis, controlling for age and number of comorbidities, revealed correlations between fatigue and all variables measured at 3- and 6-months, with the exception of self-care and time (*Table 2*).

**Table 2.** Logistic regression of PROMs follow-up variables and fatigue, adjusted for age and number of comorbidities

|  | Fatigue at 3 and 6 months |  |  |
| --- | --- | --- | --- |
|  | OR | 95% CI | p |
| **PHQ-9 Score<sup>a</sup>** | 5.05 | 1.92-13.31 | 0.001 |
| **EQ-5D Score<sup>b</sup>** | 2.35 | 1.65-3.36 | <0.001 |
| Usual Activities | 11.9 | 2.83-44.22 | 0.001 |
| Mobility | 5.39 | 1.96-14.82 | 0.001 |
| **Shortness of Breath Score<sup>c</sup>** | 1.09 | 1.04-1.14 | <0.001 |
| **PSQI Score<sup>d</sup>** | 1.71 | 1.24-2.36 | 0.001 |

Patient-Reported Outcome Measures (PROMs)
a. PHQ-9 Score, scores range from 0 – 27; 0 = *None-minimal* and 27 = *Severe*.
b. EQ-5 Score, scores range from 0 to 1; 0 = *Death* and 1 = *Perfect Health*.
c. Shortness of Breath (UCSD), scores range 0 to 120; high scores indicate greater dyspnea.
d. PSQI Score; scores range 0 to 21, higher scores indicate worse sleep quality.

### Substantial Fatigue: Hospitalization Variables

Fourteen individuals exhibited substantial fatigue at 3 months and six continued to exhibit substantial fatigue at 6 months follow-up. No patients developed new substantial fatigue between follow-up time periods. Adjusting for age, the number of comorbidities at hospitalization was associated with substantial fatigue at 3 months follow-up (OR 1.73; 95% CI 1.06-2.95; p = 0.033). No other hospitalization variables were associated with substantial fatigue.

### Substantial Fatigue: Follow-up Variables

With the exception of shortness of breath, all follow-up variables were associated with substantial fatigue at 3 months (*Table 3*). Due to the small number of cases at 6 months, we did not examine this relationship further.

**Table 3.** Predictors of Substantial Fatigue at 3 months

| **Substantial Fatigue** |  |  |  |
| --- | --- | --- | --- |
| **Hospitalization Variables** | *OR* | *95% CI* | *p* |
| Sex (Female) | 1.36 | 0.41 – 4.33 | 0.608 |
| Age | 0.74 | 0.39 – 1.44 | 0.358 |
| Number of Comorbidities | 1.73 | 1.06 – 2.95 | 0.033 |
| ICU admission | 0.77 | 0.23 – 2.44 | 0.660 |
| Ventilation | 0.28 | 0.01 – 1.60 | 0.240 |
| **3 Month Follow-Up Variables** |  |  |  |
|  | *OR* | *95% CI* | *p* |
| PHQ-9 | 1.43 | 1.23-1.78 | <0.001 |
| EQ-5D | 1.64 | 1.32-2.20 | <0.001 |
| Mobility | 2.43 | 1.47-4.33 | 0.001 |
| Usual Activities | 5.46 | 2.69-13.45 | <0.001 |
| Pain/Discomfort | 3.98 | 2.06-8.81 | <0.001 |
| Anxiety/Depression | 3.70 | 1.96-8.04 | <0.001 |
| Health Today | 0.92 | 0.87-0.96 | <0.001 |
| Shortness of Breath | 1.02 | 1.00-1.05 | 0.062 |
| PSQI | 1.43 | 1.21-1.77 | <0.001 |

## Discussion

The present study shows that long-term fatigue can persist for at least 6 months in 59.5% of patients previously hospitalized with SARS-CoV-2 infection. Within our cohort, reductions in the prevalence of fatigue and substantial fatigue were observed between 3- and 6-months.

Greater improvements were observed among those experiencing substantial fatigue, which decreased to less than half of the original prevalence by the 6-month follow-up timepoint. These findings align with existing literature reporting a decreasing trend in the proportion of individuals experiencing fatigue across weeks since acute presentation.^12,21^ Patients with more comorbidities at the time of hospitalization were also found to be more likely to exhibit fatigue at 3 and 6 months post-viral infection. Patients who did not exhibit fatigue or substantial fatigue at 3 months did not go on to exhibit fatigue or substantial fatigue at 6 months follow-up. The considerable proportion of patients continuing to exhibit symptoms of fatigue at 6 months highlights the need for further investigations into evidence-based practices that can meet the needs of those experiencing long-term fatigue after acute viral infection.

While contrary to some findings, an unexpected and subtle protective effect of age on the presence of fatigue was observed at 3 months follow-up. This finding is consistent with Subramanian et al.,^22^ who found that after adjusting for baseline covariates, age above 30 years was associated with a lower risk of reporting post-COVID symptoms. While protective effects against fatigue among those greater than 65 years of age has also been observed,^23^ other literature indicates that rates of long-COVID increase with age from about 1-2% for those in their twenties, to about 5% among those in their sixties.^24^ Indeed, post-viral fatigue syndrome or ME/CFS may affect young people (<30 years) more often. ^25^ These discrepancies may be attributed to differential reporting of symptoms according to age and other factors (e.g. those who are younger may be less accepting of feelings of disabling fatigue not previously experienced), and should be considered in future interpretations of fatigue and patient age.

Additionally, those with very severe disease presentation on admission may have had lower expectations, in relation to being back to full health in the short-term, as compared to those with less severe SARS-CoV-2 infection. Research elucidating the role age and expectation of recovery play in long-term post-viral fatigue presentation necessitates further exploration.

Pre-existing comorbidities have also previously shown associations with post-viral fatigue syndrome symptoms.^3,22^ Our research revealed, controlling for age, that the number of comorbidities a patient had was significantly associated with fatigue and its respective severity at 3 months follow-up. Interestingly, adjusted analysis of our cohort revealed dyspnea to be associated with fatigue at 3- and 6-months but was found to only be marginally associated with substantial fatigue. These findings are likely the result of the small sample size of individuals found to be experiencing substantial fatigue at follow-up timepoints. Using the same cohort of patients, Shah et al.,^13^ found dyspnea to be the most common and persistent COVID-19 recovery symptom, with 42% experiencing dyspnea at 6 months follow-up. Unexplained dyspnea, (i.e. not related to abnormalities in lung function tests or imaging), was reported in 14% and 19% of cases at 3- and 6-months, respectively.^13^ Dyspnea in post-COVID cases has been suggested to result from multiple pathophysiological mechanisms,^26^ and has also been reported in ME/CFS cases related to other causes.^27^ This symptom is also reported in dysautonomia, a common occurrence in ME/CFS.^28^

Our findings highlight the importance of further examinations into the role viral infections have in the presentation of long-term fatigue and related morbidities. While the number of individuals experiencing fatigue after viral infection is expected to decrease at follow-up timepoints, there are a subset of individuals for whom fatigue presentation and severity will persist. It is important that clinical teams remain attentive to monitoring patients for long-term fatigue after infection with SARS-CoV-2, encouraging patients to engage in practices that can aid in mitigating fatigue severity and lasting post-COVID symptoms. Our findings add to the growing body of literature illuminating the role viral infections, such as COVID-19, may have in the development of long-term symptoms and persisting fatigue.

### Limitations

This study has several limitations. Namely, our findings may be limited by the small participant sample size. As such, we were unable to examine correlates of substantial fatigue at 6 months due to limited sample size and subsequent lack of power. This prohibited examination of differences in the predictors of substantial fatigue at 3- and 6-months follow-up and prevented further investigation into those no longer exhibiting substantial fatigue at 6 months. Future research, examining changes in predictors of substantial fatigue at follow-up timepoints, are needed to clarify potential longstanding causes of substantial fatigue.

Given our inability to ascertain whether criteria for ME/CFS or post-viral fatigue were met, substantial fatigue was used as a proxy. It is therefore likely that the prevalence of post-viral fatigue syndrome found in this study is substantially higher than rates adhering to strict clinical guidelines. Likewise, gaps remain in knowledge surrounding which comorbidities may predispose individuals to greater levels of long-term fatigue and its severity after hospitalization. Several studies have identified associations between self-reported measures and fatigue related outcomes, including reductions in quality of life and cognitive impairment.^3,29^ However, there are limitations in such measures, highlighting the importance of objective outcome measures. We also note that the generalizability of study findings is limited to the sample of patients hospitalized with COVID-19 and cannot be extrapolated to those not exhibiting symptoms of COVID-19 or those who were not hospitalized.

## Conclusion

After hospitalization, a significant proportion of individuals recovering from COVID-19 will continue to experience lingering symptoms for up to 6 months. For many patients, the presentation of post-viral symptoms will manifest in increased levels of fatigue. Further investigation into the presentation of fatigue, post-COVID infection, is needed to support evidence-based care and management for individuals experiencing long-term post-viral symptoms.

## Data Availability

All data produced in the present study are available upon reasonable request to the authors

## References

1. World Health Organization. WHO Coronavirus (COVID-19) Dashboard. Published October 12, 2022. Accessed October 17, 2022. https://covid19.who.int/

2. Komaroff AL, Lipkin WI. Insights from myalgic encephalomyelitis/chronic fatigue syndrome may help unravel the pathogenesis of postacute COVID-19 syndrome. Trends Mol Med. 2021;27(9):895–906. doi:10.1016/J.MOLMED.2021.06.002

3. Ceban F, Ling S, Lui LMW, et al. Fatigue and cognitive impairment in Post-COVID-19 Syndrome: A systematic review and meta-analysis. Brain Behav Immun. 2022;101:93–135. doi:10.1016/J.BBI.2021.12.020

4. Han Q, Zheng B, Daines L, Sheikh A. Long-Term Sequelae of COVID-19: A Systematic Review and Meta-Analysis of One-Year Follow-Up Studies on Post-COVID Symptoms. Pathogens. 2022;11(2):269. doi:10.3390/PATHOGENS11020269/S1

5. Sanchez-Ramirez DC, Normand K, Yang Z, Torres-Castro R. Long-term impact of COVID-19: A systematic review of the literature and meta-analysis. Biomedicines. 2021;9(8). doi:10.3390/BIOMEDICINES9080900/S1

6. Jarrott B, Head R, Pringle KG, Lumbers ER, Martin JH. “LONG COVID”—A hypothesis for understanding the biological basis and pharmacological treatment strategy. Pharmacol Res Perspect. 2022;10(1):e00911. doi:10.1002/PRP2.911

7. National Institute for Health and Care Excellence. Myalgic Encephalomyelitis (or Encephalopathy)/Chronic Fatigue Syndrome: Diagnosis and Management. London; 2021. Accessed October 11, 2022. https://www.nice.org.uk/guidance/ng206

8. Centers for Disease Control and Prevention. Long COVID or Post-COVID Conditions. Published 2022. Accessed October 12, 2022. https://www.cdc.gov/coronavirus/2019-ncov/long-term-effects/index.html#:~:text=Estimates%20of%20the%20proportion%20of,among%20patients%20who%20were%20hospitalized

9. Zaim S, Chong JH, Sankaranarayanan V, Harky A. COVID-19 and Multiorgan Response. Curr Probl Cardiol. 2020;45(8):100618. doi:10.1016/J.CPCARDIOL.2020.100618

10. Premraj L, Kannapadi N, Briggs J, et al. Mid and long-term neurological and neuropsychiatric manifestations of post-COVID-19 syndrome: A meta-analysis. J Neurol Sci. 2022;434:120162. doi:10.1016/j.jns.2022.120162

11. Ladds E, Rushforth A, Wieringa S, et al. Persistent symptoms after Covid-19: qualitative study of 114 “long Covid” patients and draft quality principles for services. BMC Health Serv Res. 2020;20(1):1–13. doi:10.1186/S12913-020-06001-Y/TABLES/1

12. Sandler CX, Wyller VBB, Moss-Morris R, et al. Long COVID and Post-infective Fatigue Syndrome: A Review. Open Forum Infect Dis. 2021;8(10). doi:10.1093/OFID/OFAB440

13. Shah AS, Ryu MH, Hague CJ, et al. Changes in pulmonary function and patient-reported outcomes during COVID-19 recovery: a longitudinal, prospective cohort study. ERJ Open Res. 2021;7(3):00243–02021. doi:10.1183/23120541.00243-2021

14. Wong AW, Shah AS, Johnston JC, Carlsten C, Ryerson CJ. Patient-reported outcome measures after COVID-19: a prospective cohort study. European Respiratory Journal. 2020;56(5). doi:10.1183/13993003.03276-2020

15. Janssen B, Szende A. Self-Reported Population Health: An International Perspective Based on EQ-5D. 1st ed. (Szende A, Janssen B, Cabases J, eds.). Springer Dordrecht; 2014. doi:https://doi.org/10.1007/978-94-007-7596-1

16. Eakin EG, Resnikoff PM, Prewitt LM, Ries AL, Kaplan RM. Validation of a new dyspnea measure: The UCSD Shortness of Breath Questionnaire. Chest. 1998;113(3):619–624. doi:10.1378/chest.113.3.619

17. Kupferberg DH, Kaplan RM, Slymen DJ, Ries AL. Minimal clinically important difference for the UCSD Shortness of Breath Questionnaire. J Cardiopulm Rehabil. 2005;25(6):370–377. doi:10.1097/00008483-200511000-00011

18. Kroenke K, Spitzer RL, Williams JBW. The PHQ-9: validity of a brief depression severity measure. J Gen Intern Med. 2001;16(9):606–613. doi:10.1046/J.1525-1497.2001.016009606.X

19. Buysse D, Reynolds C, Monk T, Berman S, Kupfer D. The Pittsburgh Sleep Quality Index: a new instrument for the psychiatric practice and research. Psychiatry Research. 1988;28:193–213. doi:10.1016/0165-1781(89)90047-4.

20. Rockwood K, Song X, MacKnight C, et al. A global clinical measure of fitness and frailty in elderly people. CMAJ□: Canadian Medical Association Journal. 2005;173(5):489. doi:10.1503/CMAJ.050051

21. Karaarslan F, Güneri FD, Kardeş S. Long COVID: rheumatologic/musculoskeletal symptoms in hospitalized COVID-19 survivors at 3 and 6 months. Clin Rheumatol. 2022;41(1):289–296. doi:10.1007/S10067-021-05942-X/TABLES/6

22. Subramanian A, Nirantharakumar K, Hughes S, et al. Symptoms and risk factors for long COVID in non-hospitalized adults. Nature Medicine 2022 28:8. 2022;28(8):1706–1714. doi:10.1038/s41591-022-01909-w

23. Santis LVD, Pérez-Camacho I, Sobrino B, et al. Clinical and immunoserological status 12 weeks after infection with COVID-19: prospective observational study. medRxiv. Published online October 8, 2020:2020.10.06.20206060. doi:10.1101/2020.10.06.20206060

24. Bansal R, Gubbi S, Koch CA. COVID-19 and chronic fatigue syndrome: An endocrine perspective. J Clin Transl Endocrinol. 2022;27:100284. doi:10.1016/J.JCTE.2021.100284

25. Magnus P, Gunnes N, Tveito K, et al. Chronic fatigue syndrome/myalgic encephalomyelitis (CFS/ME) is associated with pandemic influenza infection, but not with an adjuvanted pandemic influenza vaccine. Vaccine. 2015;33:6173–6177. doi:10.1016/j.vaccine.2015.10.018

26. Natelson BH, Brunjes DL, Mancini D. Chronic Fatigue Syndrome and Cardiovascular Disease JACC State-of-the-Art Review. J Am Coll Cardiol. 2021;78(10):1056–1067. doi:10.1016/j.jacc.2021.06.045

27. Ravindran M, Adewuyi O, Zheng Y, et al. Dyspnea in Chronic Fatigue Syndrome (CFS): Comparison of Two Prospective Cross-Sectional Studies. Glob J Health Sci. 2013;5(2):94. doi:10.5539/GJHS.V5N2P94

28. Raj SR. Postural Tachycardia Syndrome (POTS). Circulation. 2013;127(23):2336–2342. doi:10.1161/CIRCULATIONAHA.112.144501

29. Grover S, Sahoo S, Mishra E, et al. Fatigue, perceived stigma, self-reported cognitive deficits and psychological morbidity in patients recovered from COVID-19 infection. Asian J Psychiatr. 2021;64:102815. doi:10.1016/J.AJP.2021.102815

